# Corneal neuropathy in central serous chorioretinopathy

**DOI:** 10.1101/2024.05.15.24307436

**Authors:** Jean-Louis Bourges, Bastien Leclercq, Françine Behar-Cohen

**Affiliations:** Université Paris Cité, Paris Descartes medical school, Department of ophthalmology, Cochin Hospital, Assistance Publique-Hôpitaux de Paris (APHP), France; Centre de recherche des Cordeliers, INSERM UMRS 1138 Team 17; 15 rue de l’école de médecine, 75006 Paris, France

**Keywords:** cornea, Central Serous ChorioRetinitis, confocal microscopy, nerves, dysautonomia

## Abstract

**Purpose:** Choroidal sympathetic innervation is impaired in central serous chorioretinopathy (CSCR) (1), throughout mineralocorticoid receptor.(2) We hypothesize that the CSCR condition modifies the entire eyeball innervation system. Corneal nerve shape is easily observed by in vivo confocal microscopy (IVCM).(3) We explored corneal nerves morphology in 2 groups of patients with CSCR and without (CTRL), using vivo confocal microscopy.

**Methods:** We quantified choroid thickness in both groups by OCT-EDI mode (Spectralis; Heidelberg). Patients were free from corneal disease. We explored the central, mid-peripheral, paralimbal and limbal corneal areas of patients with and without CSCR by IVCM (HRT3; Heidelberg). We proceeded with the multilayer module of acquisition and analyzed systematically the subepithelial area (SE), Bowman’s layer (BL), anterior (AS)/intermediate (IS)/deep stroma (DS) (Depth<50µm/50≤D<150µm/D≥150µm respectively), and endothelium. We semi-quantified as absent (0), rare (1) or common (2) by scoring the morphological corneal nerves abnormalities in both groups.

**Results:** We compared 15 CSCR to 11 age-matched CTRL. While abnormalities were detected sparsely (n=3; in SE, BL and A), 8 CTRL showed normal corneal nerve networks. All CSCR but one displayed nerve abnormalities. Nerve abnormalities were considered as normal, moderate, or severe in respectively 8, 3 and none CRTL versus 1, 3 and 12 CSCR. Nerves alterations were noticeably analogous to each other in all layers for CSCR. More subnormal nerve patterns were observed compared to CTRL in the 5 explored layers (p≤ 0.001).

**Conclusions:** Both thin nerve network and subepithelial plexus are altered in cornea of CSCR patients compared to CTRL, mainly across anterior layers and along straight nerve ramifications crossing the mid stroma of cornea. Nerve fiber abnormalities seems to develop in the cornea of CSCR patients in the absence of clinical corneopathy, and could serve as an early marker for the disease.

## Introduction

Most of the ocular functions are controlled by the dense innervation of tissues in the anterior and posterior segments of the eye. Autonomic innervation of the eye is ensured by parasympathetic input emerging from the III and VII cranial nerves and by sympathetic input that araise from the neck sympathetic trunk. The parasympathetic fibers of the cranial nerve III transit through the ciliary ganglion and the VII cranial nerves fibers transit through the pterygopalatine ganglion. Sensory fibers of cranial nerve V1 project to the trigeminal ganglion and to the pterygopalatine ganglion.

Denervation experiments have served to demonstrated that the three different types of nerve fibers (sympathetic, parasympathetic and sensitive) are present in nerve of the anterior segment and of the choroid and they allowed to identify neurotransmitters present in the different ocular nervous ganglia^1^. Further experiments showed that neurokinin-A is the predominant neuropeptides in the sensory fibers in the human eye although substance P and CGRP positive fibers are also released in the cornea, the trabecular meshwork, the iris, the ciliary body and the choroid.^2^.^3^

The anatomy and development of human corneal innervation was recently reviewed by Schwend et al.^4^ We will summarize breifly its main features. The cornea is densely innervated by sensory nerves but also by fibers of the autonomic nervous system, with sympathetic fibers innervating the epithelium in rodents and in humans, but with VIP-positive parasympathetic in rodents only^5^. Noradrenaline positive sympathetic fibers have been also characterized in the human corneal epithelium and vascular endothelium ^6^. The corneal nerves originate from two long ciliary nerves that extend from the ophthalmic branch of the trigeminal ganglion, branch and divide along the suprachoroidal space up to the limbus where they form the limbal plexus around the cornea. Autonomic fibers innervate the limbal vessels and sensory nerves enter into the cornea, where they lose myelin and perinerium and branch to form a dense axonal network mostly in the upper part of the anterior stroma and in the subepithelial region, referred to as the subepithelial nerve plexus (SEP). The corneal epithelium receives small nerve bundles, known as epithelial leashes. The dense network of nerves that form the sub-basal nerve plexus (SNP) are the most recognizable, their density is known to decrease with age and they are frequently affected in case of disease ^4^.

The group of Reiner and Nickla have thoroughly described the choroidal nervous system showing that sensory input from the trigeminal ganglion, sympathetic input from the superior cervical ganglion and a parasympathetic input from the pterygopalatine ganglion forms a dense and complex network ^7,8^ that is essential for controlling all choroidal functions. In humans, concentrated below the fovea, intrinsic choroidal neurons (IChNS) organized in plexi ^8–10^ contains parasympathetic, sympathetic and sensory components ^11^, suggesting that autonomous and sensory regulations ^12^ could contribute to foveal control of the blood flow and to focalization^7,8^. Recently, we have observed in both rodent and human choroid, a sensitive CGRP-positive nerve network around choroidal vessels and in non-vascularized area with connections to choroidal cells such as macrophages and mast cells, suggesting a role of this network in neurogenic inflammation.^13^

The importance of the ocular nervous system has been increasingly recognized in the pathogenesis of ocular surface diseases^14^ such as meibomian gland dysfunction^15,16^ and limbal deficiency^17^ or in dry eye^18, 19^ in which *in vivo* confocal microscopy allowed to visualize and characterize the damaged nerves^14^. In the posterior segment of the eye, no imaging method allows to analyze the choroidal nerves. But the role of the choroidal nervous system, has been demonstrated in models of dry or wet forms^20,21^ of age-related macular degeneration^22^ and more recently, choroidal neuropathy has been identified in a rodent model of pachychoroid induced by the overexpression of the mineralocorticoid receptor^13^.

Pachychoroid is an imaging choroidal vascular phenotype with dilated choroidal veins and effacement of the choriocapillaris, eventually associated with detectable alterations of the overlying retinal pigment epithelium^23–25^. Pachychoroid predisposes to a range of diseases, including central serous chorioretinopathy (CSCR) but the exact mechanism of pachychoroid remains unclear, related to genetic predisposition^26^, veinous overload^27^, scleral rigidity ^28^ and to an abnormal neural control of the choroidal blood flow. Indeed, we have hypothesized that the vascular changes, observed in pachychoroid could originate, at least in part from a choroidal neuropathy^13^. The systemic dysfunction of the autonomous nervous system measured in patients with CSCR supports this hypothesis, such as low heart rate variability ^29,30^, abnormal pupillometric response ^31^ and the correlation between choroidal thickness and heart rate variability in CSCR patients^32^. But whether choroidal nerves show neuropathic signs in CSCR is unknown.

As the choroidal nervous system cannot be imaged by the current imaging techniques, we have investigated the corneal nerves in patients with CSCR using *in vivo* confocal microscopy and have compared them to unaffected control subjects.

## Methods

### Design of the study

We prospectively enrolled consecutive CSCR patients and control volunteers with a normal choroid who consented to our monocentric unmasked observational comparative study from May 2023 to February 2024. All patients gave their informed consent. The national IRB approved the protocol under Unique Protocol ID# 2022-A02713-40 and ClinicalTrials.gov registration.

### Population and criteria

We analyzed both eyes of a same patient and pooled our observations under the same patient’s ID. We included consecutive patients above 18 years who met the following inclusion criteria.

The CSCR group consisted of consecutive patients with chronic CSCR complicated by epitheliopathy, pachychoroid, subfoveolar choroidal thickness (SFCT) >390µm, features and history of subretinal fluid and area of alteration of the pigment retinal epithelium of blue autofluorescence superior to three disk diameters or multifocal.

The control group (CTRL) enrolled volunteers with subfoveal choroidal thickness ≤350µm, and cleared from any retinal or corneal disease.

### Data collection

We quantified choroid thickness in both groups by OCT-EDI mode (Spectralis; Heidelberg) prior to proceed to the *in-vivo* confocal microscopy (IVCM; HRT3; Heidelberg) corneal exploration. We proceeded with the multilayer module of acquisition of the IVCM. Briefly, we tested corneal sensitivity prior to anesthetize nociception with chlorhydrate oxybuprocaïne eye drops repeated twice in 10 min. Then we docked IVMC toward the central area of cornea. We set acquisition to the most superficial layer of epithelium and proceeded longitudinally toward corneal periphery by acquiring both snapshot frames and multilayer sequences, to encompass the greater area possible. We explored the central first, followed by mid-peripheral, paralimbal, and endothelial corneal areas. We analyzed the corneal nerve network systematically across 5 corneal layers: Bowman’s layer (BL), subepithelial area (SE), anterior (AS)/mid (MS)/deep stroma (DS). At least, one sequential acquisition was performed in the central subepithelial layers in depth, completed with the acquisition of more than 500 analyzable pictures for each patient.

Under masked condition for the CSCR or CTRL group, we semi-quantified abnormalities in both groups. Two experienced operators, one subspecialized in corneology (JLB) and the other subspecialized in vitreoretinopathy (FFBC), proceeding during independent masked rounds of reading, scoring nerve abnormalities as absent (0), rare (1), frequent (2) or omnipresent (3) by comparing the morphological corneal nerves to a same gold standard. (20, 21) We screened morphological abnormalities observed in the population of the study, chasing typical reccurent patterns described in Figure 1.

**Figure 1:**
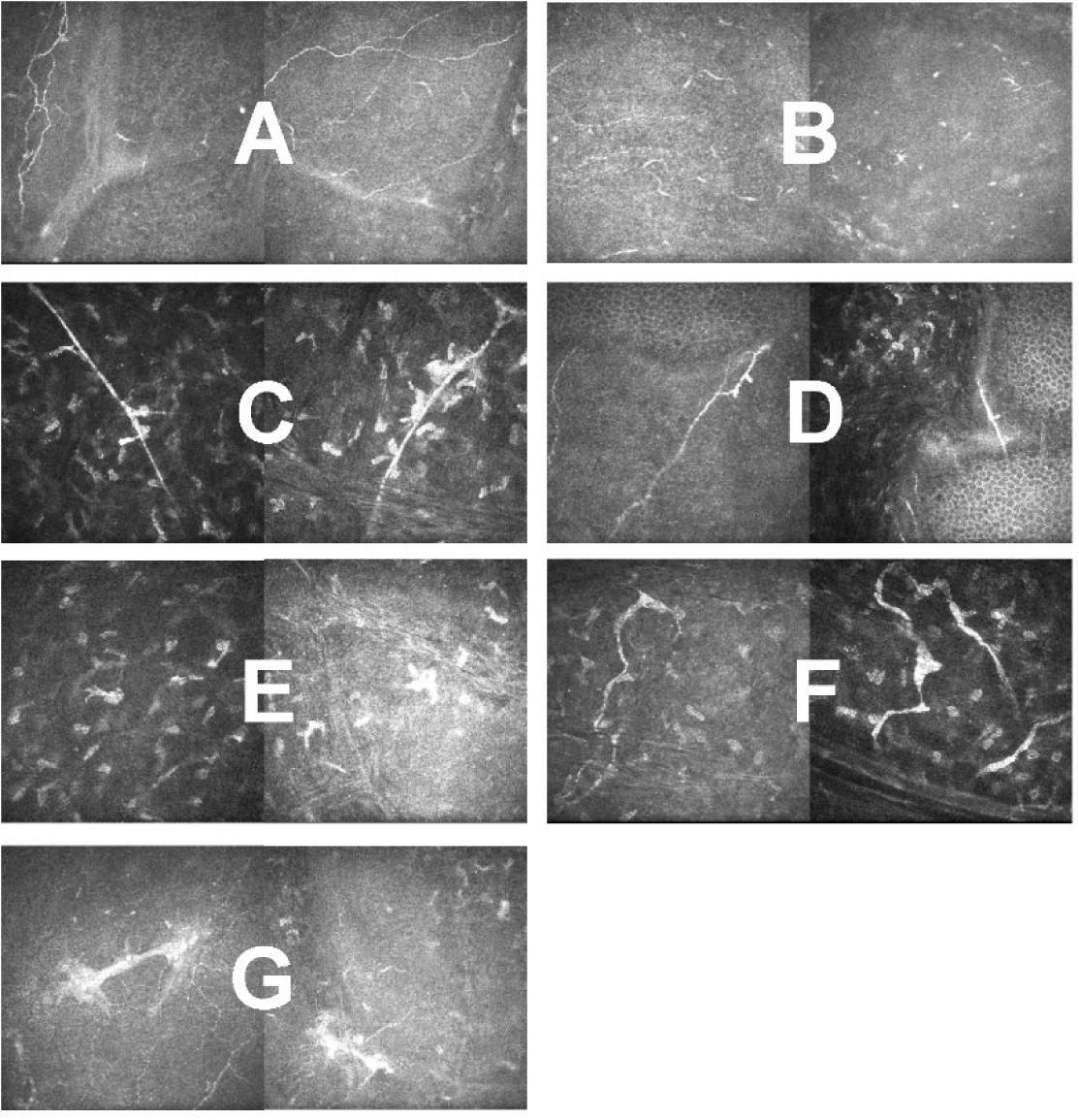
Abnormal entities of the neural network of cornea and observed in our population of study. A: terminal nerve loops and crossing fibers; B: rarified terminal nerve network; C:cell adhesion to the nerve fiber edges; D: Bulbar excrescences along nerve fibers; E: activation of ectopique dendritique cells; F: thickening and irregularity of of the anterior stromal network of nerve fibers; G: sub basal large neuroma.

First, we tested the concordance of scores between observers. Then, we analyzed layer by layer the attributed scores to compare CSCR to CTRL groups. We qualified the corneal nerve network of each patient using the grading system described in Tables and figures.

Table 1. We arbitrarily assumed that the corneal nerve network was normal in the absence of affected layers, or if some rare nerve abnormalities were observed in only 1 or 2 layers, being agreed by both readings. Finally, we qualified the whole corneal nerve network according to the global density.

**Table 1:** Grading system to qualify the corneal nerve network across the 5 corneal layers analyzed.

| Score attributed to a single layer | # layers concerned | Interpretation proposed |
| --- | --- | --- |
| 0 or 1 | $n \leq 2$ | Normal corneal nerves |
| 2 | $n < 2$ | Moderate corneal nerves abnormality |
| 2 | $n \geq 2$ | Severe corneal nerves abnormality |
| 3 | $n \neq 0$ | Severe corneal nerves abnormality |

**Table 2:**
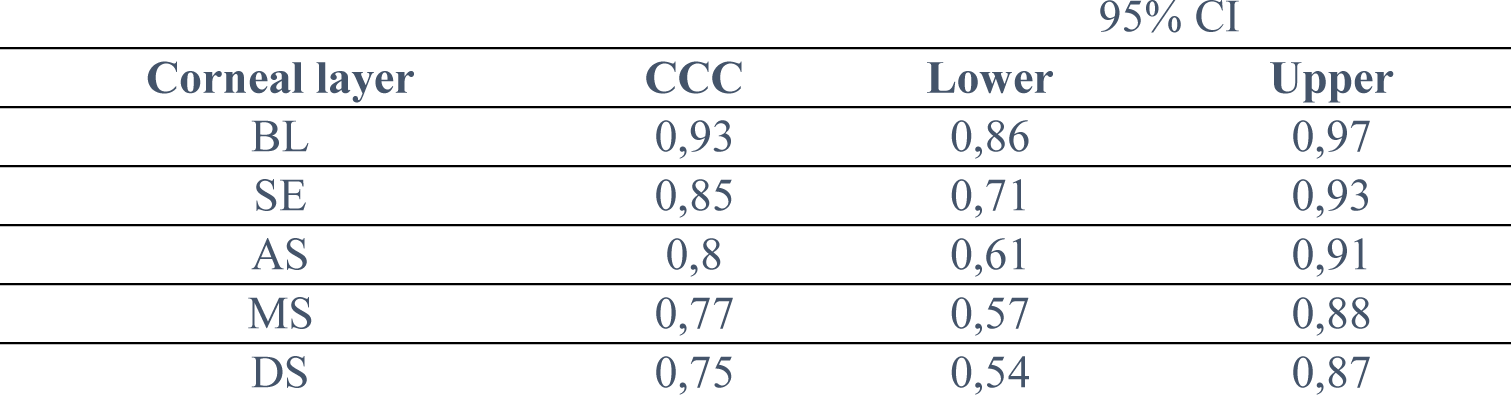
Estimated Correlation of Concordance (CCC) for nerve abnormality scores across corneal layers between the two readers.

### Statistical analysis

We extracted the data by tabulation in an Excel sheet (Microsoft Office Standard version 2016). We proceeded with the help of Jamovi 2023 and modules ad hoc (v.2.3; the Jamovi project, https://www.jamovi.org) to analyze the information collected from the above-described scale. To evaluate observations of the clinical condition proposed and due to the observational pilot nature of this study, we did not proceed to formal sample size calculation nor applied methods for multiplicity. Descriptive statistics were computed for all items. We tested the score agreement between the two operators (JLB and FBC) by calculating the concordance correlation coefficient (CCC; upper-lower at 95%CI) for each scored layer measures the agreement. We considered CCC values < 0.90 as Poor, 0.90 - 0.95 as Moderate, 0.95 - 0.99 as Substantial and >0.99 as Almost perfect. We carried out the comparison of the two groups by testing variance egality with the Levene test (p<0,05). We applied the t-test when variances in case of variance egality, or the Mann-Whitney U test for non-parametric data and Kruskal-Wallis when appropriate. The statistical significance was defined as p<0.05 and taking confidence level as 95%.

## Results

We initially screened 15 CSCR and 15 CTRL. Among them, we could not obtain interpretable IVCM pictures from 4 CTRL patients: 2 did not show for examination and 2 had lid spastic reflexes incompatible with the IVCM examination. Finally, we enrolled 15 CSCR patients and 11 CTRL. The mean age was 49.5 years (from 28 to 83 y) for CTRL and 57.2 years (from 37 to 85 yo) for CSCR (ns; p=0,12). All corneas were avascular, exempt from detectable opacity at slit-lamp examination. All corneas had full sensitivity to light touch.

The mean subfoveal thickness was measured at 497 µm±111 µm [451-543µm] and 262 µm± 80 µm [226-297µm] respectively in the CTRL and the CSCR groups (p<0.0001 Man-Whitney U test). The correlation of nerve abnormality scores between reader rated from good to excellent for all corneal layers except for the endothelium for which the agreement remained moderate (In the CTRL group, 8 individuals showed entirely normal corneal nerves. Moderate abnormalities were detected in 3/11 IVMC, located anteriorly in SE (n=3), BL (n=2), AS (n=3) and MS (n=1). No more than 3 layers were involved. We rated 3 corneas as moderately abnormal, but none were severely altered.

In the CSCR group, A single individual was considered as normal, however with mild nerve fiber alterations. Others shown moderate (n=2) to severe (n=12) nerves disorders. The difference was high versus CTRL (p≤ 0.001).

Superficial and mid layers were preferentially involved compared to deep ones or versus CTRL (p≤ 0.005). At least, two layers or more displayed subnormal patterns. In the Bowman’s layer, we observed twisted and rarified nerve fibers, occasionally forming unusual large loops. We also observed multiple area of hypertrophic patterns within the subepithelial nerve plexus, as well as waved nerve edges evoking possible neuroma features at both anterior and mid stromal nerve layer’s detriment (Figure 2). A single CSCR individual was considered as normal with mild nerve fiber alterations however, while others shown moderate (n=2) to severe (n=12) disorders (Figure 3).

**Figure 2:**
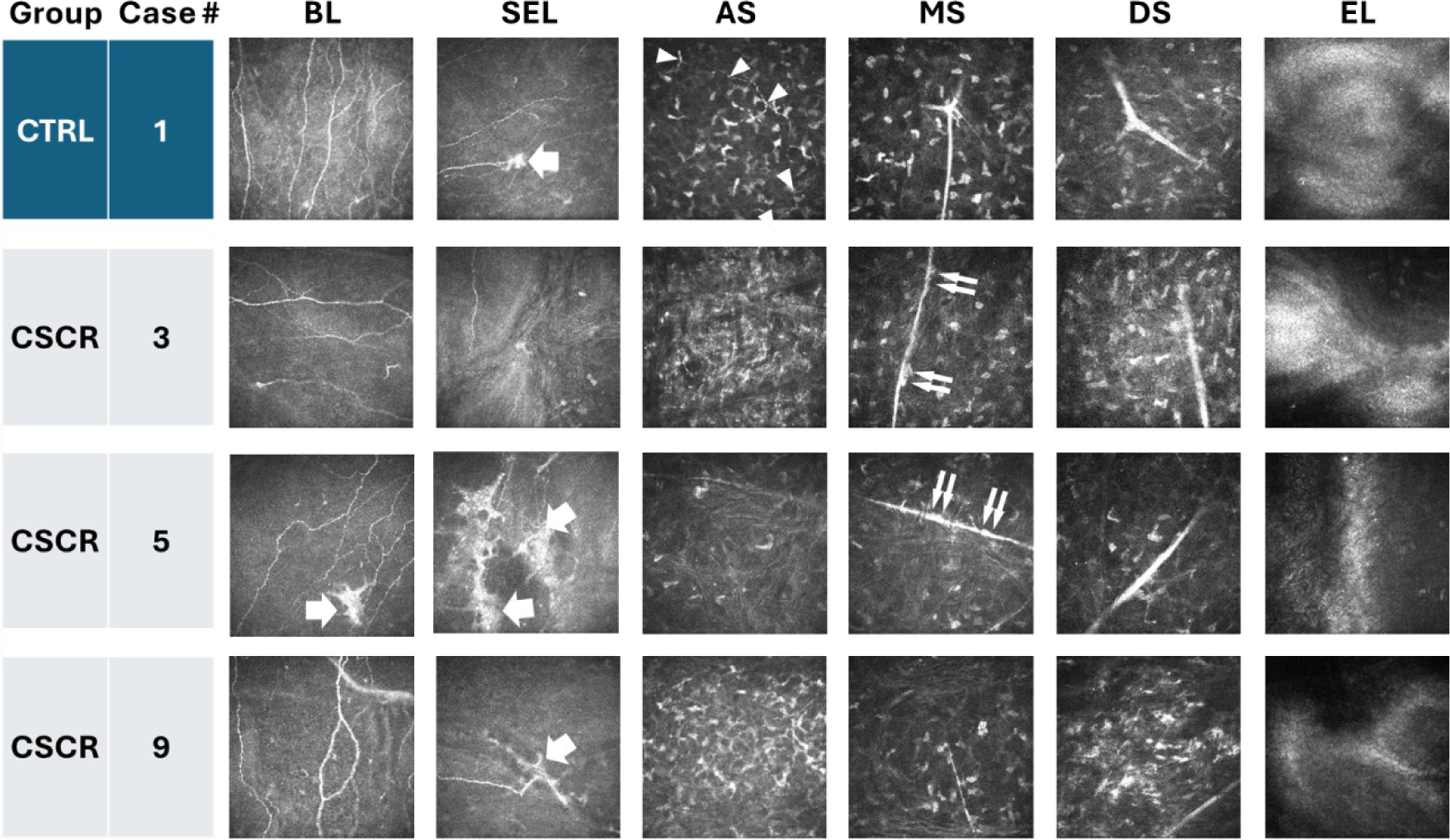
In vivo confocal microscopy (IVMC) frames from controls (CTRL) and central serous chorioretinopathy (CSCR). Examples of pictures obtained from Bowman’s layer (BL), subepithelial area (SEL), corneal stroma at different depth (anterior =AS, mid=MS; Deep=DS), and endothelium (EL). Nerve fibers striated densely BL in CTRL’s eyes, whereas we observed a weaker network in CSCR (BL and SEL; case 3, case 9) with unusually superficial hypertrophic structures (BL; case 5) and wide loops (BL; case 9). The subepithelial nerve plexus (thick arrows) appeared either enlarged and granulated or elongated (SEL; case 5) with fuzzy edges (SEL; case 5 and 9). The fine and scalloped nerve network of the anterior stroma (arrowheads) was commonly rarefied or undetectable, within a variably hypo-reflective matrix (AS column). Crossing the MS, the long and normally straight nerves (CTRL, MS; case 1) displayed swelled and moniliform edges (double arrows, cases 3 and 5).

**Figure 3:**
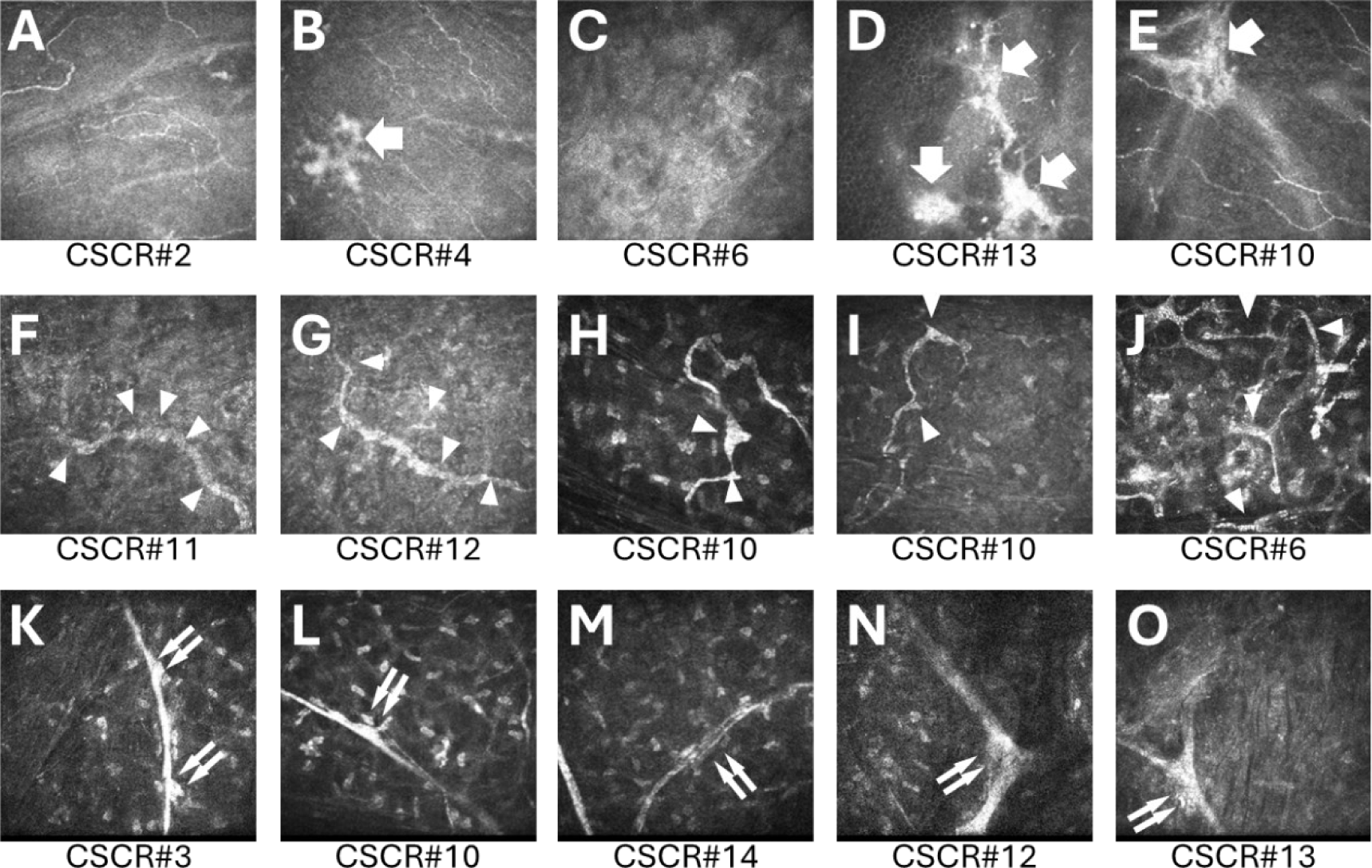
In vivo confocal microscopy (IVMC) examples of characteristic nerve fiber’s alterations observed from CSCR eyes. At the level of Bowman’s layer (BL; A, B, C) and subepithelial area (SEL; C, D, E), we observe a decreased density of nerve fibers, with hypertrophic neuroma-like nerve plexi (arrows). The anterior stroma (AS, F to J) featured thickened ant irregular nerve branches (arrowheads). The deep nerve structures were swelled and blistered with iso-reflective lumps (K,L; double arrows) while some others seemed of less and double edged reflectivity (M). At the intersection of the large and deep corneal nerve troncs (N, O), we observed occasionally some irregular hyper-reflectivity (N,O; double arrows).

Noticeably, nerves were altered within all explored layers significantly more in CSCR compared to CTRL (p≤ 0.001, Figure 4). While nerve fiber seemed normally dense in CTRL, almost half CSCR (n=7/15) revealed marked and diffuse rarefaction. Logically and deprived from nerve fibers, the endothelial scores seemed not differ between groups (p=0,56). Superficial and mid corneal layers were preferentially involved rather than deep ones and compared to CTRL (Mann-Whitney; p≤ 0.005). Endothelial patterns again remained normal in both groups.

**Figure 4:**
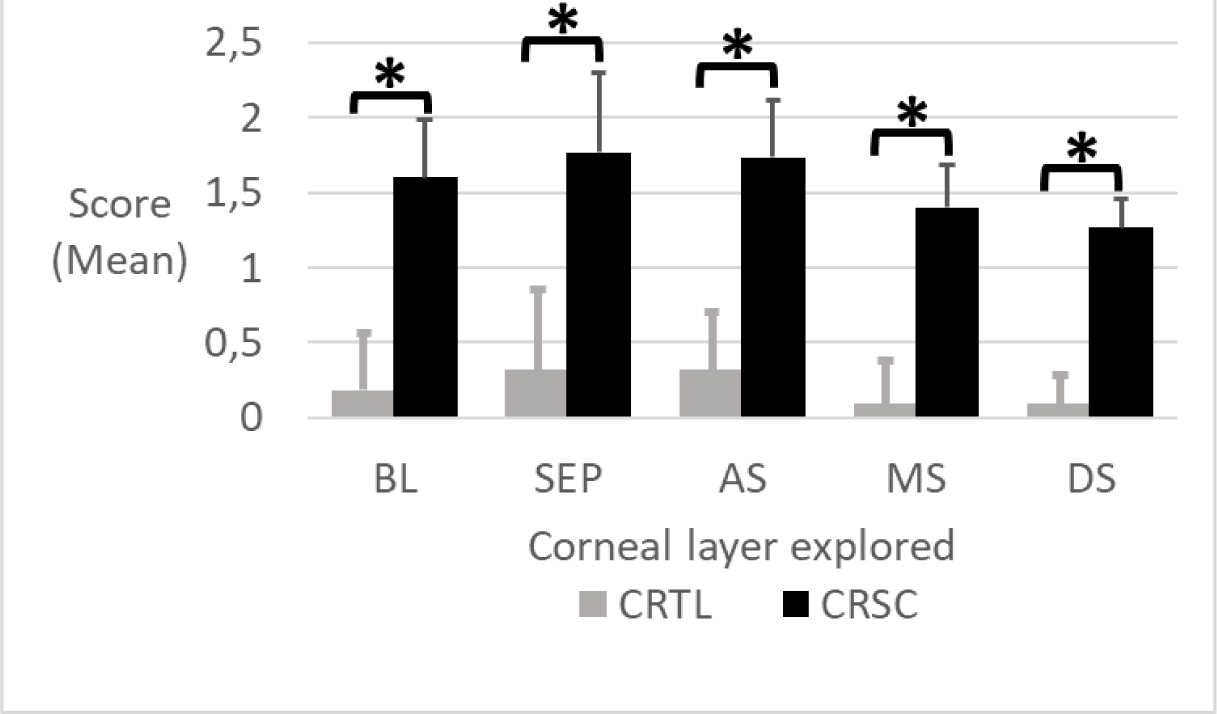
Mean scores of corneal nerve abnormality observed by vivo confocal microscopy (IVMC) from controls (CTRL) and central serous chorioretinopathy (CSCR). *: significative difference, Kruskal-Wallis test, p value<0.001.

## Discussion

Because of its transparency and easy access, the cornea is an ideal tissue to analyze in a non-invasive manner, the peripheral sensory nerves, that are affected frequently in ocular but also in systemic diseases, although patients might present minimal or no symptoms.

In this study, we have performed a systematic analysis of the corneal nerves in all corneal layers in patients with severe forms of CSCR and pachychoroid features and have identified morphological signs of corneal neuropathy in these patients. Abnormalities of nerves were clearly identified by two independent graders in the different corneal layers and not only were they observed more frequently (14/15, 93%) in eyes with CSCR as compared to the unaffected controls (3/11, 27%) but the lesions were more pronounced in the CSCR group. In the basal/ subbasal layer, there was a rarefaction in 50% of the CSCR patients’cornea with no difference in the nerve tortuosity, that is frequently observed in this age group of patients33. Microneuroma, that could correspond to disorganized and aggregated Schwann cells and fibroblasts surrounding degenerated axon endings34, were a frequent finding in this layer, although we could observe them in one of the cornea in the control group. Such microneuroma are a common but non specific indicator of abnormal regeneration of axons. Other signs of nerve pathology35 were observed such as coiling, looping and twisting of tortuous stromal nerves, stubs of degenerated stromal nerves and signs of remodeling such as sprouting. Dendritic cells activation could be observed in 11/15 (75%) of the CRSC cornea. These nerves abnormalities were not associated with any clinically detectable corneal pathology and no characterized ocular surface pathology. Whether patients with CSCR have a higher rate of dry eye disease has not been studied and the examined patients did not suffer from dry eye symptoms. It is thus unlikely that the observed neuropathy could be secondary to dry eye disease in this group of patients.

The mechanism of corneal neuropathy in CSCR could be related to a more general alteration of the peripheral nerves. In a recent paper, we have hypothesized that the choroidal hyperreflective organized structures, typically observed around the dilatated vessels in pachychoroid patients, could correspond to abnormal choroidal nerves ^13^, similar to the abnormal structure of the choroidal nerves we could image in rats overexpressing the mineralocorticoid receptors and a pachychoroid-like phenotype13,36. The observation of abnormal nerves in the cornea of CSCR patients, together with the functional signs of dysautonomia in these patients ^29,30^ ^31^ ^32^ is highly suggestive that autonomic neuropathy contributes to the disease. Similar findings have been repeatedly reported in patients suffering from autonomic diabetic neuropathy37 and in which the cornea has served as a sensor of the severity and the risk of cardiovascular events. In diabetic type 1, corneal nerve fibre metrics predicted incident cardiovascular and cerebrovascular events38, though mechanisms involving neuroimmune inflammation related to increased sympathetic activity within the vascular wall. In diabetic patients, corneal confocal imaging has been considered as a rapid, non-invasive, but also highly sensitive and specific for the diagnostic of autonomic diabetic neuropathy.^39^.

Evidence of corneal nerve abnormalities have been also described in patients familial dysautonomia, that could lead up neurotrophic keratitis40. In CSCR, whether corneal nerve abnormalities are early signs and whether they could be detected in patient with uncomplicated acute CSCR is currently under evaluation. Its prognosis value also should be further evaluated. The fact that nerves, outside from the choroid show signs of neuropathy favors the hypothesis that choroidal neuropathy is not secondary to CSCR but might contribute to the disease pathogenesis. Other features associated with CSCR such as sleep apnea41, ^42^, hypertension, arrythmia43 or post traumatic stress disorders44 have been also associated with autonomous nerve dysfunction, which could constitute an unifying link between CSCR and other favoring factors including systemic or extraocular corticotherapy, through the induction of an imbalance of corticoid receptors45 and MR-induced choroidal neuropathy13.

The mechanisms of autonomis nervous system dysfunction and of corneal neuropathy in CSCR remains to be deciphered. It could be related to a combination of genetic and environmental / exogenous factors. In particular, low C4B CNV (copy variation number) was strongly associated with severe forms of CSCR46 and C4 plays a major role in monocytic-induced synapse pruning47 that is a mechanism required for neural remodeling. In favor of this hypothesis, we found that systemic lipocalin 248,49 and pentraxin 350, both induced by corticoids but significantly reduced in CSCR patients, play important role in neural remodelling ^51^. Genetic and exogenous factors might concur to a progressive peripheral neuropathy in patients with chronic forms of CSCR, affecting more specifically, but not exclusively the autonomous nervous system.

Limitations of the study include the fact that our analysis is qualitative but there are no automatic method to quantify all the corneal abnormalities observed with confocal microscopy. This weakness is counterbalanced by the blind scorring of two observers and the imaging of all the cornea surface of patients, which tend to reduce biais. Another limitation is that we did not assess in depth ocular surface functional parameters such as lachrymal function in our patients and controls but this should be performed in a larger cohort to analyse if CSCR is associated with any infraclinical ocular surface disease. Only patients with severe choroidopathy and epitheliopathy were included and further ongoing study in prospectively analyzing cornel nerves in patients with acute CSCR and with pachychoroid.

In conclusion, this is to our knowledge, the first observation that patients with chronic CSCR present signs of corneal neuropathy, which support our previous observation that choroidal neuropathy could contribute to the disease pathogenesis. Further studies on larger cohorst are needed to confirm these findings.

In the CTRL group, 8 individuals showed entirely normal corneal nerves. Moderate abnormalities were detected in 3/11 IVMC, located anteriorly in SE (n=3), BL (n=2), AS (n=3) and MS (n=1). No more than 3 layers were involved. We rated 3 corneas as moderately abnormal, but none were severely altered.

In the CSCR group, A single individual was considered as normal, however with mild nerve fiber alterations. Others shown moderate (n=2) to severe (n=12) nerves disorders. The difference was high versus CTRL (p≤ 0.001).

Superficial and mid layers were preferentially involved compared to deep ones or versus CTRL (p≤ 0.005). At least, two layers or more displayed subnormal patterns. In the Bowman’s layer, we observed twisted and rarified nerve fibers, occasionally forming unusual large loops. We also observed multiple area of hypertrophic patterns within the subepithelial nerve plexus, as well as waved nerve edges evoking possible neuroma features at both anterior and mid stromal nerve layer’s detriment (Figure 2). A single CSCR individual was considered as normal with mild nerve fiber alterations however, while others shown moderate (n=2) to severe (n=12) disorders (Figure 3).

Noticeably, nerves were altered within all explored layers significantly more in CSCR compared to CTRL (p≤ 0.001, Figure 4). While nerve fiber seemed normally dense in CTRL, almost half CSCR (n=7/15) revealed marked and diffuse rarefaction. Logically and deprived from nerve fibers, the endothelial scores seemed not differ between groups (p=0,56). Superficial and mid corneal layers were preferentially involved rather than deep ones and compared to CTRL (Mann-Whitney; p≤ 0.005). Endothelial patterns again remained normal in both groups.

## Discussion

Because of its transparency and easy access, the cornea is an ideal tissue to analyze in a non-invasive manner, the peripheral sensory nerves, that are affected frequently in ocular but also in systemic diseases, although patients might present minimal or no symptoms.

In this study, we have performed a systematic analysis of the corneal nerves in all corneal layers in patients with severe forms of CSCR and pachychoroid features and have identified morphological signs of corneal neuropathy in these patients. Abnormalities of nerves were clearly identified by two independent graders in the different corneal layers and not only were they observed more frequently (14/15, 93%) in eyes with CSCR as compared to the unaffected controls (3/11, 27%) but the lesions were more pronounced in the CSCR group. In the basal/ subbasal layer, there was a rarefaction in 50% of the CSCR patients’cornea with no difference in the nerve tortuosity, that is frequently observed in this age group of patients^33^. Microneuroma, that could correspond to disorganized and aggregated Schwann cells and fibroblasts surrounding degenerated axon endings^34^, were a frequent finding in this layer, although we could observe them in one of the cornea in the control group. Such microneuroma are a common but non specific indicator of abnormal regeneration of axons. Other signs of nerve pathology^35^ were observed such as coiling, looping and twisting of tortuous stromal nerves, stubs of degenerated stromal nerves and signs of remodeling such as sprouting. Dendritic cells activation could be observed in 11/15 (75%) of the CRSC cornea. These nerves abnormalities were not associated with any clinically detectable corneal pathology and no characterized ocular surface pathology. Whether patients with CSCR have a higher rate of dry eye disease has not been studied and the examined patients did not suffer from dry eye symptoms. It is thus unlikely that the observed neuropathy could be secondary to dry eye disease in this group of patients.

The mechanism of corneal neuropathy in CSCR could be related to a more general alteration of the peripheral nerves. In a recent paper, we have hypothesized that the choroidal hyperreflective organized structures, typically observed around the dilatated vessels in pachychoroid patients, could correspond to abnormal choroidal nerves ^13^, similar to the abnormal structure of the choroidal nerves we could image in rats overexpressing the mineralocorticoid receptors and a pachychoroid-like phenotype^13,36^. The observation of abnormal nerves in the cornea of CSCR patients, together with the functional signs of dysautonomia in these patients ^29,30^ ^31^ ^32^ is highly suggestive that autonomic neuropathy contributes to the disease. Similar findings have been repeatedly reported in patients suffering from autonomic diabetic neuropathy^37^ and in which the cornea has served as a sensor of the severity and the risk of cardiovascular events. In diabetic type 1, corneal nerve fibre metrics predicted incident cardiovascular and cerebrovascular events^38^, though mechanisms involving neuroimmune inflammation related to increased sympathetic activity within the vascular wall. In diabetic patients, corneal confocal imaging has been considered as a rapid, non-invasive, but also highly sensitive and specific for the diagnostic of autonomic diabetic neuropathy.^39^.

Evidence of corneal nerve abnormalities have been also described in patients familial dysautonomia, that could lead up neurotrophic keratitis^40^. In CSCR, whether corneal nerve abnormalities are early signs and whether they could be detected in patient with uncomplicated acute CSCR is currently under evaluation. Its prognosis value also should be further evaluated. The fact that nerves, outside from the choroid show signs of neuropathy favors the hypothesis that choroidal neuropathy is not secondary to CSCR but might contribute to the disease pathogenesis. Other features associated with CSCR such as sleep apnea^41, 42^, hypertension, arrythmia^43^ or post traumatic stress disorders^44^ have been also associated with autonomous nerve dysfunction, which could constitute an unifying link between CSCR and other favoring factors including systemic or extraocular corticotherapy, through the induction of an imbalance of corticoid receptors^45^ and MR-induced choroidal neuropathy^13^.

The mechanisms of autonomis nervous system dysfunction and of corneal neuropathy in CSCR remains to be deciphered. It could be related to a combination of genetic and environmental / exogenous factors. In particular, low C4B CNV (copy variation number) was strongly associated with severe forms of CSCR^46^ and C4 plays a major role in monocytic-induced synapse pruning^47^ that is a mechanism required for neural remodeling. In favor of this hypothesis, we found that systemic lipocalin 2^48,49^ and pentraxin 3^50^, both induced by corticoids but significantly reduced in CSCR patients, play important role in neural remodelling ^51^. Genetic and exogenous factors might concur to a progressive peripheral neuropathy in patients with chronic forms of CSCR, affecting more specifically, but not exclusively the autonomous nervous system.

Limitations of the study include the fact that our analysis is qualitative but there are no automatic method to quantify all the corneal abnormalities observed with confocal microscopy. This weakness is counterbalanced by the blind scorring of two observers and the imaging of all the cornea surface of patients, which tend to reduce biais. Another limitation is that we did not assess in depth ocular surface functional parameters such as lachrymal function in our patients and controls but this should be performed in a larger cohort to analyse if CSCR is associated with any infraclinical ocular surface disease. Only patients with severe choroidopathy and epitheliopathy were included and further ongoing study in prospectively analyzing cornel nerves in patients with acute CSCR and with pachychoroid.

In conclusion, this is to our knowledge, the first observation that patients with chronic CSCR present signs of corneal neuropathy, which support our previous observation that choroidal neuropathy could contribute to the disease pathogenesis. Further studies on larger cohorst are needed to confirm these findings.

## Data Availability

All data produced in the present study are available upon reasonable request to the authors

## Acknowledgments

The authors gratefully thank Dr Neil Lagali for his support and advice.

